# Explainable Suicide Phenotyping from Initial Psychiatric Evaluation Notes Using Reasoning Large Language Models

**DOI:** 10.1101/2025.03.27.25324783

**Authors:** Zehan Li, Wanjing Wang, Lokesh Shahani, Rodrigo M Vieira, Salih Selek, Jair Soares, Hongfang Liu, Ming Huang

**Affiliations:** McWilliams School of Biomedical Informatics, The University of Texas Health Science Center at Houston, TX, USA; Faillace Department of Psychiatry & Behavioral Sciences, McGovern Medical School, The University of Texas Health Science at Houston, Houston, TX, USA

## Abstract

Clinical phenotyping is the process of extracting patient’s observable symptoms and traits to better understand their disease condition. Suicide phenotyping focuses more on behavioral and cognitive characteristics, such as suicide ideation, attempt, and self-injury, to identify suicide risks and improve interventions. In this study, we leveraged the latest reasoning models, namely 4o, o1, and o3-mini, to perform note-level multi-label classification and reasoning generation tasks using previously annotated psychiatric evaluation notes from a safety-net psychiatric inpatient hospital in Harris County, Texas. Compared with the previously finetuned GPT-3.5 model, the out-of-box reasoning models prompted with in-context learning achieved comparable and better performance, with the highest accuracy of 0.94 and F1 of 0.90. We implemented novel clinical justification generation from these models on the traditional classification tasks. This finding marked a promising direction for performing clinical phenotyping that is interpretable and actionable using smaller, efficient reasoning models.

## Introduction

### Suicide Phenotyping

A fundamental challenge in psychiatric research and clinical practice is to accurately identify patients with specific psychopathological traits for risk assessment, early intervention, and treatment planning^1^. In biomedicine, this process is defined as clinical phenotyping, which refers to the systematic identification of observable characteristics—ranging from physiological to behavioral traits—that help define disease states and patient subpopulations^2^. Traditionally, clinical phenotyping has been instrumental in domains like genetics, oncology, and cardiology, where well-defined phenotypic profiles aid in diagnostics and therapeutic decisions^3^. With the widespread adoption of Electronic Health Records (EHRs) in clinical practice, EHR-based phenotyping has emerged as a method to identify clinical conditions, diseases, or patient characteristics by extracting structured and unstructured data from medical records. However, in psychiatry, where conditions often lack clear biological markers, phenotyping presents unique challenges due to the subjective and heterogeneous nature of mental disorders^4,5^.

It is well established that the risk of suicide is particularly high among individuals with mental health disorders^6,7^. The risk of suicide is significantly elevated in persons with a history of self-harm or previous suicide attempts (SA). Additionally, thoughts of suicide or death, known as suicide ideation (SI), often precede SA, making early identification and intervention critical^6^. Studies indicate that psychiatric disorders and substance abuse are involved in approximately 90% of all cases of suicide, highlighting the critical need for effective mental health care and intervention strategie^8^. Accordingly, being able to accurately identifying and categorizing the multifaceted nature of suicidality in individual with psychiatric disorders is imperative for developing targeted intervention, efficient resource allocation, and effective treatment plans that can save lives and improve clinical care.

### Challenges in Suicide Data Collection

Suicide phenotyping, as a specific clinical phenotyping, involves identifying behavioral and cognitive markers for suicide events and related factors from existing data^9,10^. While data on suicide deaths is available through the public health registries, such as National Death Index, comprehensive information on the continuum of suicidality remains limited^11^. Traditional methods, such as structured psychometric surveys, are hindered by recall bias and stigma^12^. Among individuals with serious mental health conditions, psychiatric episodes and involuntary admissions further prevent the collection of high-quality structured data. More recently, the increased adoption of EHRs presents an opportunity to leverage large scale clinical data for suicide phenotyping and related observational study. However, such studies remain limited due to challenges in psychiatric data accessibility^13^. For example, in our previous study, only 3% of SI and 19% of SA cases in clinical notes had corresponding diagnostic codes^14^. Consequently, this limitation restricts the scope and effectiveness of EHR-based observational and epidemiological studies. Observed or disclosed suicidal tendencies are typically recorded by providers in the patient’s clinical narrative, regardless of whether these observations are linked to diagnostic codes and/or included in the billing reports. Unstructured clinical narratives, particularly inpatient psychiatric evaluation notes, contain rich textual data on suicidality, capturing details on mental health status, life stressors, and behavioral risk factors^15^. As a result, these narratives represent a critical yet underutilized resource for suicide phenotyping, offering significant potential for downstream analysis.

### Clinical NLP and Related Works

Natural Language Processing (NLP) presents a promising approach to extracting suicide phenotypes from psychiatric evaluation notes, addressing the reporting gap left by structured billing data^16,17^. Previously, single-class classification approaches, such as predicting the presence or absence of SA, have been widely used. Recent work has increasingly focused on multi-label classification for suicidality detection using NLP^18–21^. These studies can be categorized into two groups based on classification methods: multiple binary classification and single multi-label classification. The multiple binary classification approach treats each suicidality category separately, where models independently predict the presence or absence of SI or SA. For instance, one study developed a sentence level binary classification task based on RoBERTa models and achieved F1 score of 0.83 for either SI or SA and 0.78 in differentiating SI from SA, showcasing a reliable approach for finetuning pretrained transformer models with an additional annotated psychiatric corpus^20^. In contrast, single multi-label classification models account for the co-occurrence of multiple suicidal events and related factors, better capturing the complexity of real-world psychiatric presentations. In fact, patients admitted to psychiatric services often present with SI in conjunction with a past or recent SA or Non-Suicidal Self-Injury (NSSI). In our previous study, we found significant overlapping among annotated suicide labels with 45% of the notes having more than 2 labels^14^. Using multi-label neural network approach, our finetuned model resulted in a 4% improvement in performance compared to RoBERTa trained with binary relevance method, achieving a micro-averaged F1 score of 0.81 ± 0.01^21^.

### Reasoning Language Models

Reasoning Language Models (RLMs) are advanced generative AI models designed to handle complex reasoning tasks that surpass standard text generation^22,23^. Models like DeepSeek R1 and GPT-o1 exemplify this approach, using reasoning tokens to break down prompts into smaller steps, evaluate solutions, and dynamically adjust responses^24,25^. These models extend traditional large language models (LLMs) by emulating human cognitive processes, enabling them to perform logical inference, solve multi-step problems, and engage in nuanced reasoning^26^. Specifically, after generating reasoning tokens, the model produces an answer as visible completion tokens and discards the reasoning tokens from its context. Utilizing such structured reasoning techniques, such as Chain-of-Thought prompting, RLMs can explicitly outline their reasoning processes, improving transparency and interpretability^23^. Consequently, they excel in tasks requiring mathematical reasoning, logical deduction, planning, and strategic thinking, not merely providing answers but also clearly explaining the reasoning behind them.

### Study Objectives

With the recent advancement in LLMs, we aim to explore the application of reasoning LLMs for psychiatric suicide phenotyping. Specifically, we investigate their ability to perform multi-label classification of suicide phenotypes from initial psychiatric evaluation notes (IPEs) and to generate clinically relevant reasoning for error analysis. We also aim to compare classification performance between fine-tuned traditional LLM and in-context learned RLMs. By leveraging knowledge in generative AI and in-context learning, we aim to assess whether reasoning models can improve AI-driven phenotyping performance while offering interpretable, clinically meaningful insights into suicide events and related factors.

## Methods

In the following sections, we outline the steps to perform multi-label classification to identify suicidal events and factors in Initial Psychiatric Evaluation (IPE) notes with RLMs. We started with collection, annotation, and analysis of the study dataset, including characteristics and distribution of labels. We then describe the RLMs and the multi-label classification approach employed. Additionally, we detail the experimental setup and conclude by evaluating model performance at both the overall model and individual label levels (**Figure 1**). This study was approved by the Institutional Review Board (IRB: HSC-SBMI-17-0354) at the University of Texas Health Science Center at Houston.

**Figure 1.**
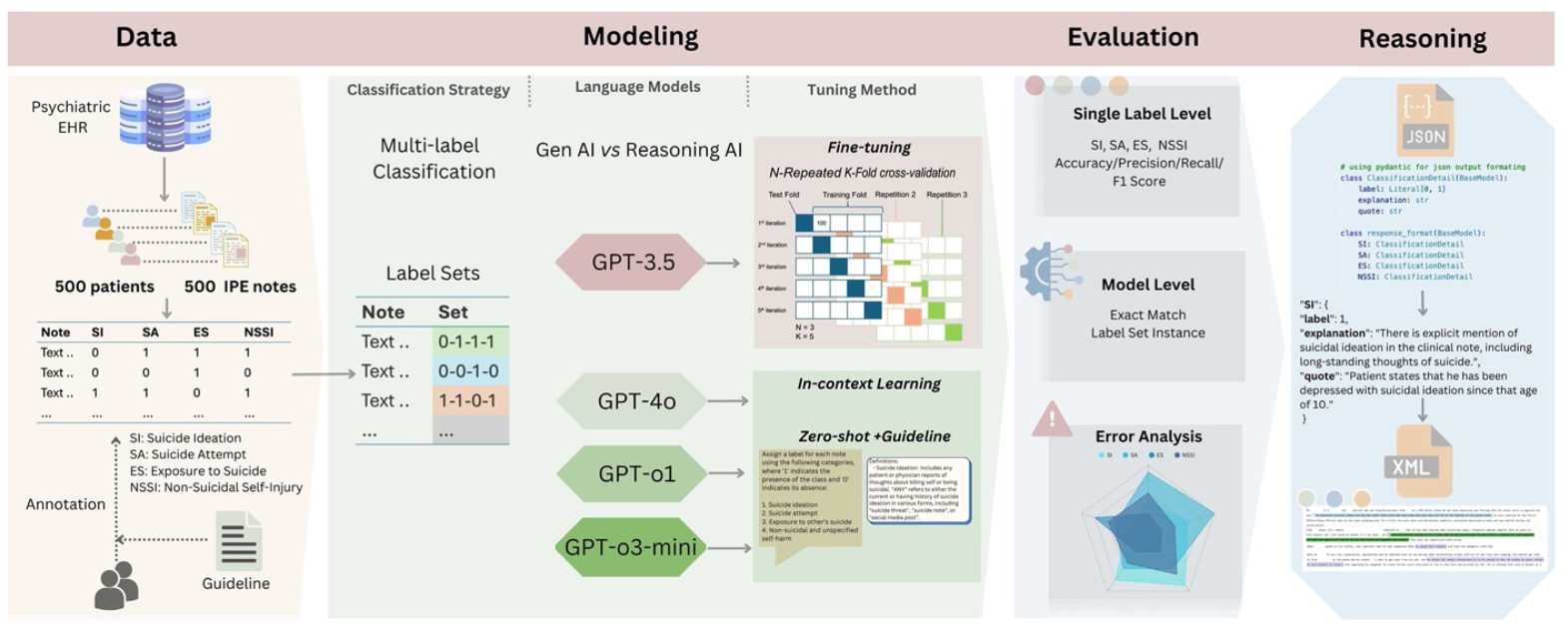
Graphical Abstract on data processing, language modeling, automatic evaluation, and classification reasoning

### Psychiatric inpatient dataset

For this study, we adopted a previously annotated dataset of 500 IPE notes from the EHR database of a safety-net psychiatric inpatient hospital, Harris County Psychiatric Center (HCPC)^14^. These notes were labeled with four suicide-related categories: “Suicidal Ideation” (SI), “Suicide Attempts” (SA), “Exposure to Suicide” (ES), and “Non-Suicidal Self-Injury” (NSSI). SI involves thoughts or plans about suicide or killing oneself, whereas SA denotes actual suicidal behaviors to end one’s life. ES refers to experiences involving exposure to another person’s suicide, and NSSI describes deliberate self-injury without suicidal intent. The IAA includes 0.98 for SI, 1.00 for SA, 0.98 for ES, and 0.90 for NSSI, indicating near-perfect agreement (0.81–1.00)^27^.

The distribution of each label is shown in Figure 2. The 500 notes were assigned with a total of 675 labels, specifically 294 SI, 265 SA, 22 ES, and 94 NSSI. Of the 500 notes, 103 are free from any suicide mentions. A total of 172 notes (34.4%) contained only one label, with the majority being SI (N=96) or SA (N=62), while smaller counts included NSSI (N=11) and ES (N=3). Conversely, 225 notes (45%) contained more than one label, with the most common combination being SI and SA (N=178, 35.6%). Furthermore, a subset of 45 notes (9%) contained three labels, while 4 notes (0.8%) contained all four labels (SI, SA, ES, and NSSI). This distribution reflects the high acuity and complexity of suicidal behaviors in the inpatient psychiatric population at HCPC, highlighting the coexisting nature of suicidal events and related factors.

**Figure 2.**
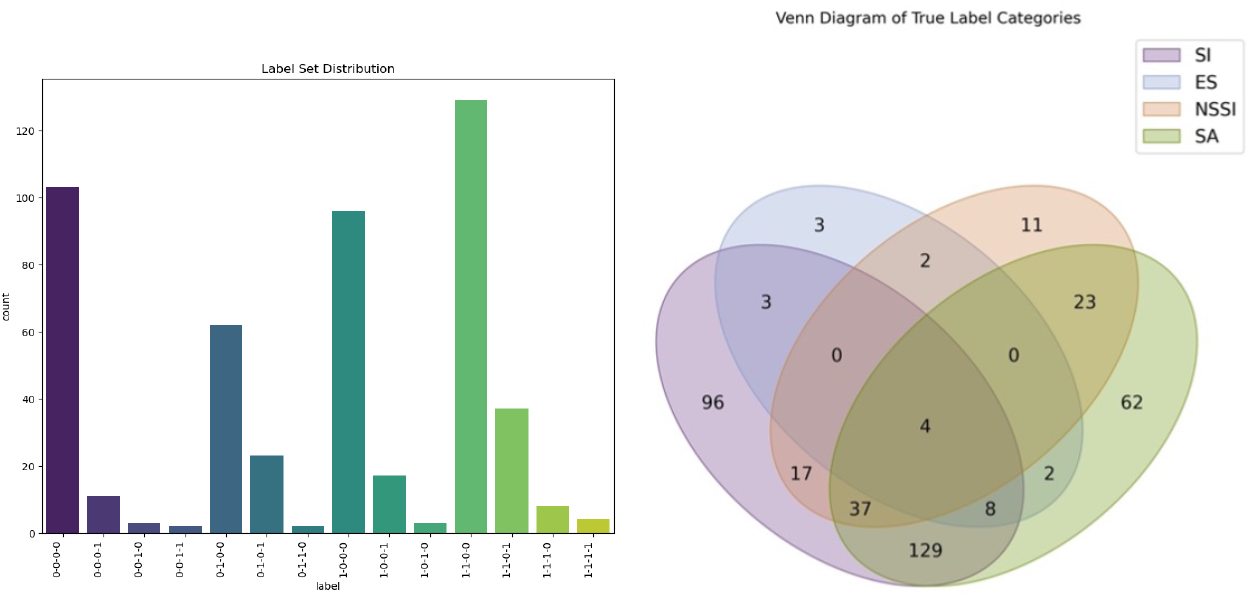
Label set distribution showing coexisting suicidal events and related factors. Label sequence: SI-SA-ES-NSSI

### Reasoning Language Models

In this study, we leveraged the latest RLM, namely GPT-4o, GPT-o1, and GPT-o3-mini from OpenAI to perform multi-label classification to identify multiple suicidal events and risk factors within the IPE notes. GPT-4o (“Omni”) is a multimodal language model capable of processing and generating text, images, and audio. GPT-o1 is OpenAI’s first language model emphasizing advanced reasoning abilities. It employs reinforcement learning techniques to process queries step-by-step, akin to human reasoning. This approach enables GPT-o1 to tackle complex tasks, particularly in coding, mathematics, and scientific reasoning. GPT-o3-mini is a compact and efficient model optimized for rapid responses and precise reasoning. GPT-o3-mini excels in tasks requiring quick and accurate reasoning, making it suitable for applications where speed and cost-efficiency are crucial.

### Multi-label Classification

To identify the multiple suicidal events and factors at the document level, we employed a multi-label classification approach by leveraging RLMs. Multi-label classification is a classification paradigm in which each instance can simultaneously belong to multiple labels or categories, rather than being limited to a single category. This approach effectively captures the complexity of real-world scenarios, where data points often possess multiple attributes or characteristics that cannot be adequately described by a single label. It is particularly suitable for tasks where assigning multiple labels provides a more accurate and comprehensive representation of the data.

Multi-label classification enables the simultaneous identification of multiple labels that are not mutually exclusive. For our multi-label classification task, we designed a prompt based on the annotation guideline instructing the RLMs to output a set of four binary codes (0 or 1) in JSON format. Each binary digit indicates the presence (1) or absence (0) of one of the four labels, for example, {“SI”: 1, “SA”: 1, “ES”: 0, “NSSI”: 1}.

### Reasoning Generation

We performed reasoning generation tasks with these RLMs to generate structured, logical explanations through explicit reasoning about the identification of multiple suicidal events and risk factors. On top of the classification prompt, we added three additional instructions: (1) Refer to the guideline above and clinical knowledge to generate comprehensive explanation to justify each label classification; (2) Quote from the text to support reasoning; (3) Follow the output format strictly to make sure every output has all the properties. The included RLMs have new “*response_format”* parameter which was defined with two class functions. Figure 3 shows an example of reasoning generation using NSSI.

**Figure 3.**
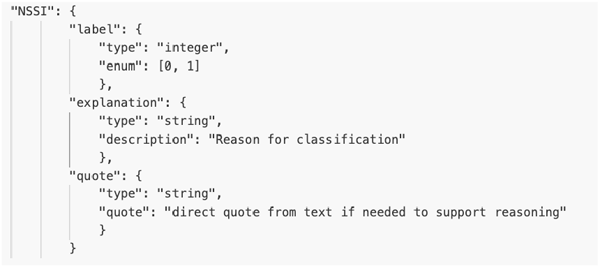
A prompt example of reasoning generation using NSSI.

Another new parameter exclusive for the RLMs is “*reasoning_effort”*, which gives the model guidance on how many reasoning tokens it should generate before creating a response to the prompt. For this study, we set the “*reasoning_effort”* to “high” for all RLMs, favoring more complete reasoning at the cost of more tokens generated and slower responses.

### Model training and Evaluation

We implemented multi-label classification using RLMs to identify multiple suicidal events and related factors at the document level. These RLMs were accessed via the OpenAI API hosted on Microsoft Azure Studio, a HIPAA-compliant cloud computing platform. We deployed zero-shot learning using GPT-4o, GPT-o1, and GPT-o3-mini to classify suicidal events and related factors in IPE notes. A structured prompt with was developed and tested for zero-shot learning for multi-label classification, instructing these GPT models to generate multi-label predictions and associated reasoning in JSON format for 500 IPE notes. These reasoning models were compared with a previously best-performing fine-tuned GPT model, GPT-3.5-Tune^28^. The GPT-3.5 model was previously fine-tuned by feeding pairs of IPE notes as input and sets of four labels as output. To evaluate model performance, we calculated accuracy, precision, recall, and F1 scores at both the label and model levels. Specifically, each metrics was computed individually for each label to assess label-specific performance. Additionally, overall accuracy and micro-average precision, recall, and F1 scores were calculated to summarize model performance.

## Result

### Model Level

Figure 4 illustrates the exact match accuracy across the four models. GPT-o3-mini achieved the highest exact match accuracy of 0.79, followed closely by GPT-o1 at 0.77, GPT-3.5-Tune at 0.78, and GPT-4o at 0.70, highlighting strong performance of GPT-o3-mini in multi-label classification with exact match.

**Figure 4.**
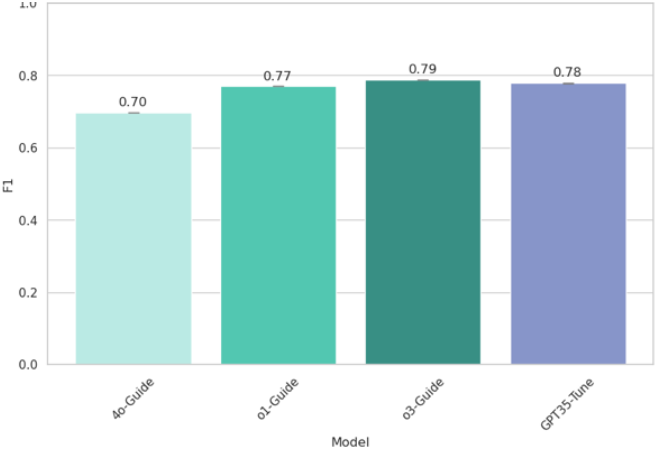
Model performance in terms of exact match F1 score

In terms of micro-averaged metrics of single labels (**Figure 5**), GPT-o3-mini and GPT-3.5-Tune exhibited the best performance with an accuracy of 0.94 and an F1 score of 0.90. GPT-o3-mini achieved the highest precision at 0.92, comparable to GPT-3.5-Tune which also showed a precision of 0.91. GPT-4o showed a higher recall of 0.94, but lower precision and F1 scores compared to the remaining models, suggesting trade-offs in model performance characteristics.

**Figure 5.**
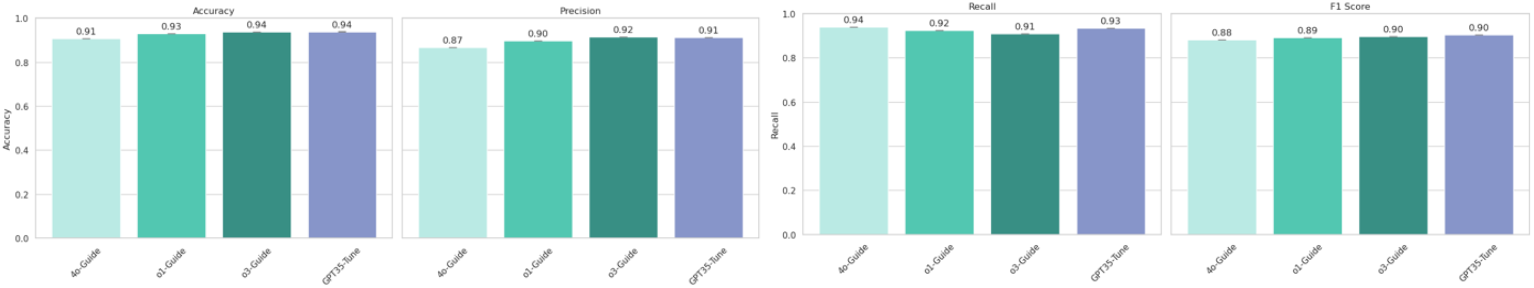
Model performance in terms of micro-averaged accuracy, precision, recall, and F1 scores

### Single Label Level

The evaluation performance for each of the four suicidal labels (SI, SA, ES, and NSSI) is detailed in **Figure 6**. Accuracy remained consistently high across all models and labels, with the highest variability observed in precision, recall, and F1 scores. For SI and SA labels, GPT-o3-mini and GPT-3.5-Tune consistently outperformed GPT-4o and GPT-o1, with GPT-o3-mini achieving an F1 score of 0.91 for SI and 0.90 for SA. Notably, GPT-4o displayed the lowest performance for these labels, particularly impacting its overall lower exact match accuracy.

**Figure 6.**
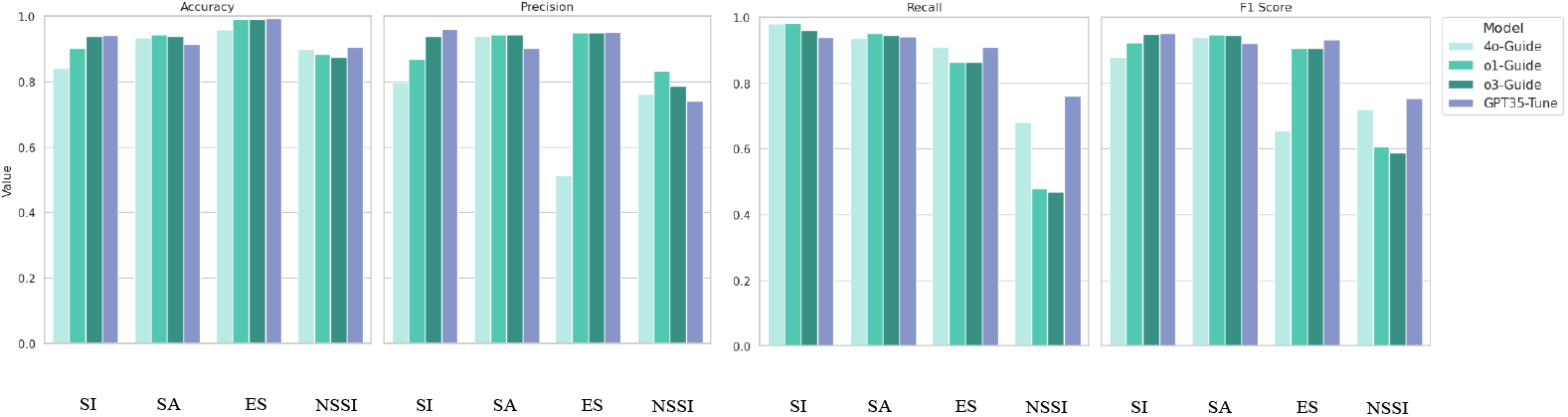
Model performance for each label

The ES label demonstrated significant label imbalance challenges. GPT-o3-mini showed a notably higher F1 score (0.65) than GPT-4o (0.40), GPT-o1 (0.50), and GPT-3.5 (0.60), indicating robustness in handling imbalanced labels. For the NSSI label, GPT-3.5-Tune achieved the highest F1 score (0.73), followed closely by GPT-o3-mini (0.72), outperforming GPT-o1 (0.65) and GPT-4o (0.58). This highlights the efficiency of GPT-o3-mini and GPT-3.5-Tune in managing less common but clinically significant labels such as NSSI. Overall, the smaller, more efficient reasoning model GPT-o3-mini demonstrated superior or comparable performance across most suicidal labels, emphasizing its robustness and suitability for clinical suicide phenotyping from psychiatric evaluation notes.

### Error Analysis

Given that GPT-o3-mini demonstrated comparable or superior performance across individual labels and overall classification, we selected it for further examination of clinical reasoning generation. To analyze output errors, we selected one note from each of the top 10 most frequent misclassified label sets (Table 1). These cases included both false positives (FP) and false negatives (FN). We found that most misclassifications involved a single label (Cases 1–8), with fewer instances of multiple-label misclassification (Cases 9 and 10). We recognized a pattern of frequent misclassification on NSSI, a total of 40 FN of the 70 selected instances. This issue was particularly prevalent when SA was also present, suggesting that the model struggled to differentiate between SA and NSSI when both appeared in the same note (Cases 1, 3, and 9). Additionally, three instances exhibited a higher degree of confusion, where SA and NSSI were misclassified for each other (Case 10). This difficulty was further reflected in the single-label performance, where NSSI had the lowest F1 score (0.59), indicating a persistent challenge for the model in distinguishing between these two conceptually similar phenotypes.

**Table 1.** Error analysis on GPT-o3-mini. Label sequence: SI-SA-ES-NSSI. Rank by frequency.

| Case | True_labels | Predict_labels | Frequency | Error Type |
| --- | --- | --- | --- | --- |
| 1 | 1-1-0-1 | 1-1-0-0 | 22 | FN on NSSI |
| 2 | 1-0-0-0 | 0-0-0-0 | 9 | FN on SI |
| 3 | 0-1-0-1 | 0-1-0-0 | 9 | FN on NSSI |
| 4 | 1-0-0-1 | 1-0-0-0 | 5 | FN on NSSI |
| 5 | 1-0-0-0 | 1-1-0-0 | 5 | FP on SA |
| 6 | 0-1-0-0 | 1-1-0-0 | 5 | FN on SI |
| 7 | 0-0-0-0 | 1-0-0-0 | 4 | FP on SI |
| 8 | 0-0-0-0 | 0-1-0-0 | 4 | FP on SA |
| 9 | 0-1-0-1 | 1-1-0-0 | 4 | FP on SI, FN on NSSI |
| 10 | 1-1-0-0 | 1-0-0-1 | 3 | FN on SA, FP on NSSI |

### Reasoning

For the above error instances, we promoted the GPT-o3-mini to generate clinical reasoning to justify the classification output as well as decision evidence from the notes. We selected one instance to demonstrate the structured reasoning output capability for error analysis.

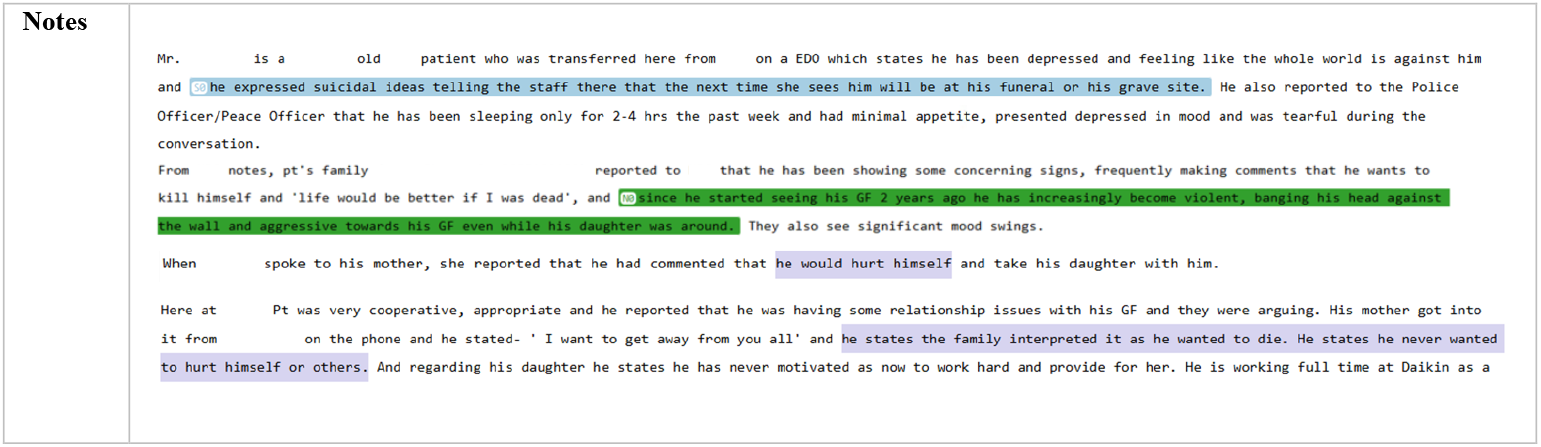

**Table 2.** Example using one error case on NSSI.

| Label | True | Predict | Reasoning | Decision Evidence |
| --- | --- | --- | --- | --- |
| SI | 1 | 1 | Multiple clinical observations and reports indicate clear suicidal ideation. The note documents that Mr. Colbert expressed suicidal ideas during his transfer from NPC, with comments interpreted by staff and family members, even though his later self-report denies such thoughts. Given that physician observations take precedence, his earlier statement is decisive. | "he expressed suicidal ideas telling the staff there that the next time she sees him will be at his funeral or his grave site" |
| SA | 0 | 0 | There is no evidence in the narrative of an actual suicide attempt or behavior intended to cause self-death. The documentation only reflects ideation without described self-destructive actions. |  |
| ES | 0 | 0 | The note does not mention incidents involving suicides of others or external suicide events. All references focus on Mr. Colbert's own ideation. |  |
| NSSI | 1 | 0 | While there is mention of behaviors such as banging his head against the wall, these are reported in the context of overall agitation and violence rather than deliberate self-injury without suicidal intent. Furthermore, the patient explicitly denies wanting to hurt himself. | "banging his head against the wall" |

According to the annotation guidelines and prompt, NSSI includes behaviors driven by agitation, such as “banging head,”. In this case, the model correctly identified relevant evidence but over-interpreted the scope of NSSI by following a stricter definition than what was defined in the prompt. Additionally, conflicting information within the note further complicated the classification. Our prompt included a General Rule to guide evidence prioritization: *“Give precedence to a physician’s observations and judgments over reports from the patient’s family/friends, followed by the patient’s self-report*.*”*. However, the reasoning output did not adhere to this hierarchy, instead prioritizing the patient’s denial over his mother’s report. This deviation highlights a limitation in the model’s ability to consistently follow structured reasoning rules, suggesting that future improvements should focus on reinforcing evidence prioritization through few-shot prompting.

## Discussion

### In-context Learned Reasoning Model

Our results demonstrate that the reasoning models achieved performance comparable to, and in exact match analysis, even surpassed the fine-tuned GPT-3.5 model. This highlights the efficacy of advanced reasoning architectures in handling complex psychiatric text without requiring domain-specific fine-tuning. Traditionally, fine-tuning has been the state-of-the-art approach for clinical NLP tasks, as it allows models to adapt specifically to medical terminology and domain-specific expressions. Fine-tuned models have been widely adopted in clinical applications such as named entity recognition (NER), relation extraction (RE), and other phenotyping tasks, as they provide highly specialized performance improvements over general-purpose language models. Fine-tuning offers key advantages by allowing models to learn the specialized clinical expression, reduces hallucinations and enhances performance for tasks requiring high specificity. However, fine-tuning is computationally expensive, often requiring large amounts of labeled data and extensive hyperparameter tuning. Additionally, model updates require retraining on new data, making it less generalizable for rapidly evolving clinical settings.

In contrast, in-context learning with advanced reasoning models presents a paradigm shift for clinical NLP. These models can process highly structured prompts, enabling dynamic adaptation to specific clinical tasks without retraining and large annotated dataset. With the latest reasoning architectures, which support structured outputs and accommodate long-context inputs (up to 200,000 tokens), complex clinical guidelines and even structured ontology can be integrated into prompts. This allows models to infer nuanced suicidality phenotypes, align outputs with psychiatric best practices, and perform sophisticated reasoning over extensive patient histories without domain-specific fine-tuning.

Additionally, pretrained GPT models include default content filters designed to restrict harmful content input and output. While we removed the default filter and applied customized one for this study to process psychiatric notes containing sensitive language, their existence suggests that the models have been pretrained on data distributions that inherently classify suicide related content. This may contribute to their strong baseline performance, even without fine-tuning. Furthermore, these models likely benefit from built-in classifiers that recognize self-harm-related language, reinforcing their ability to handle psychiatric text with a degree of reliability straight out of the box.

Moreover, we observed that model performance was not influenced by label distribution. For instance, although the study corpus contained only 22 cases of ES, performance on this label was comparable to more abundant categories such as SI (294) and SA (265). This suggests that reasoning models are inherently better at handling small datasets and demonstrate superior comprehension of rare events. Unlike traditional machine learning approaches, which often struggle with class imbalance and require resampling techniques, reasoning models appear to mitigate this challenge through their ability to generalize effectively from context and prompted knowledge. This is particularly beneficial in clinical settings, where certain psychiatric phenotypes may be infrequently documented but remain critical for patient risk assessment and intervention.

We argue that by leveraging in-context learning, reasoning models can now be directly applied to clinical NLP tasks with minimal overhead. Freed from the constraints of manual parameter tuning and annotation, researchers and clinicians can focus on integrating validated clinical knowledge into well-structured prompts. This significantly reduces the barriers to implementation, making it possible to deploy AI-driven solutions for suicide risk assessment, mental health documentation analysis, and other psychiatric applications with greater efficiency. While fine-tuning remains valuable for certain high-stakes applications requiring high sensitivity or specificity, the flexibility and scalability of in-context reasoning models suggest a shift in the field toward prompt-based adaptation as a computationally efficient and clinically viable alternative.

### Insights into Confusion

One notable challenge in classification was the confusion between SA and NSSI. While these two categories are mutually exclusive based on suicidal intent, they often share overlapping behavioral and contextual features in psychiatric notes. Additionally, all four phenotypes are temporally agnostic, meaning that both historical and current instances of SA are classified as positive cases in the same way. As a result, chronic NSSI and recent SA can coexist within a single note, complicating accurate classification. The models’ occasional misclassification of SA and NSSI suggests that further refinement in contextual understanding is necessary, potentially through enhanced prompt design or incorporating additional structured knowledge about suicide risk behaviors. One potential solution is to refine the prompt by explicitly instructing the model to label both SA and NSSI when suicidal intent is ambiguous, ensuring a more nuanced classification approach that reduces false negatives on NSSI.

### Limitations

This study has several limitations. First, the latest reasoning models removed several key parameters, such as top_p and temperature, which traditionally allow users to control the rigidity and variability of the model’s output. The absence of these parameters reduces the ability to control for response consistency, leading to undesired fluctuations in classification outcomes. In our experiments, we observed cases where the same psychiatric note, when reprocessed multiple times, yielded different classification results and reasoning paths. This inconsistency raises concerns about the reproducibility of model outputs, particularly in high-stakes clinical applications where reliability is critical. Future studies should consider running repeated experiments to quantify variability and establish confidence intervals, ensuring more stable model performance in real-world deployment. Second, reasoning generation was primarily used for error analysis rather than being systematically applied to all psychiatric notes. While reasoning outputs provided valuable insights into model classification decision, their effectiveness as clinical justifications were not formally evaluated. Future research should focus on assessing not only the automatic classification performance but also the quality and clinical validity of generated justifications through experienced psychiatrists. Such human evaluations will determine whether AI-generated reasoning is appropriate to aid clinical decision-making, improve trust in AI-driven phenotyping, and support real-world psychiatric practice.

## Conclusion

This study demonstrates the effectiveness of leveraging advanced reasoning models for suicide phenotyping in psychiatric evaluation notes. By employing in-context learning with models such as 4o, o1, and o3-mini, we achieved performance comparable to, and in some cases surpassing, fine-tuned GPT-3.5. This highlights the potential of reasoning models to process complex psychiatric text without the need for extensive domain-specific fine-tuning, making them a scalable and efficient alternative for clinical NLP tasks. Furthermore, our implementation of reasoning-based clinical justification generation represents a novel approach to enhancing interpretability in AI-driven phenotyping. Unlike traditional classification models that provide only categorical predictions, reasoning models offer explanations for their classifications, making their outputs interpretable for clinicians. Additionally, our findings suggest that reasoning models are particularly advantageous in settings with small or imbalanced datasets. This underscores their robustness in clinical applications where data scarcity is a common challenge. Despite these promising results, future studies should conduct repeated experiments to quantify variability and to conduct expert evaluation on the reasoning content. Overall, this study marks a significant step forward in applying advanced reasoning architectures to clinical phenotyping. By reducing the need for costly fine-tuning and leveraging structured prompts for real-time adaptation, reasoning models offer a computationally efficient and clinically viable approach to suicide risk assessment and broader psychiatric NLP applications.

## Data Availability

All data produced in the present study are available upon reasonable request to the authors and UTHealth Houston

https://www.medrxiv.org/content/10.1101/2025.01.13.25320491v1

